# Serum IgG antibodies induced by the synthetic carbohydrate-based conjugate vaccine candidate SF2a-TT15 against *Shigella flexneri* 2a cross-react with the heterologous lipopolysaccharide of *Shigella flexneri* 6

**DOI:** 10.64898/2026.05.05.26352385

**Authors:** Valeria Asato, Shiri Meron-Sudai, Anya Bialik, Sophy Goren, Shubham Mathur, Jonas Ståhle, Göran Widmalm, Armelle Phalipon, Laurence A. Mulard, Dani Cohen

## Abstract

**Background:** *Shigella flexneri* 2a (SF2a) and 6 (SF6) are two of the most common *S. flexneri* serotypes. They have distant O-specific polysaccharide (O-SP) structures. Previous studies showed no cross-reactivity or cross-protection between the two serotypes in a guinea pig model of infection. However, partial cross-reactivity and cross-protection were reported in humans immunized with a SF2a lattice-type conjugate vaccine candidate comprising the chemically detoxified lipopolysaccharide (LPS) attached to recombinant *Pseudomonas aeruginosa* Exoprotein A (rEPA).

**Objectives:** This study aimed at deciphering the possible cross-reactivity with heterologous SF6 strains of antibodies induced in humans by SF2a-TT15, a sun-type SF2a conjugate vaccine candidate featuring a non-*O*-acetylated synthetic oligosaccharide (OS) as surrogate of the detoxified LPS. Special focus was on the impact of the O-SP non-stoichiometric *O*-acetylation on cross-reactivity.

**Methods:** Serum IgG antibody titers to LPSs from SF6 strains harboring different degrees of O-SP *O*-acetylation, and from *Escherichia coli* O147 (EC147) which shares an identical but non-*O*-acetylated O-SP with SF6, were measured by ELISA in 63 serum samples of volunteers receiving 2 µg and 10 µg OS doses of SF2a-TT15 or placebo in the frame of a phase I clinical study. Antibody in-lymphocyte-supernatants (ALS), avidity, and serum bactericidal activity (SBA) were measured in a subset of volunteers.

**Results:** SF2a-TT15 induced cross-reacting IgG antibodies to all SF6 LPSs and EC147 LPS. A ≥4-fold rise in anti-SF6 IgG titers was more frequent with the 10 µg dose than with 2 µg (50% vs 22%, p=0.045). Cross-reactivity rate was higher with the low *O*-acetylated SF6 O-SP than with the high *O*-acetylated one (50% versus 21%, p<0.05). Anti-SF6 responses correlated with homologous anti-SF2a LPS responses. Similar cross-reactivity was detected in ALS samples at day 7 after vaccination. Cross-reacting antibodies were partially functional against the heterologous SF6 parental strains, as shown by bactericidal activity and increased avidity.

**Conclusions:** SF2a-TT15 induces stronger SF6 cross-reactive IgG responses than the previously tested detoxified *O*-acetylated SF2a LPS–rEPA conjugate. While both serotypes are included in most multivalent *Shigella* vaccine candidates, cross-reactivity and cross-protection between SF2a and SF6 could enhance the immunogenicity and efficacy of a *Shigella* multivalent vaccine candidate, particularly in infants in low- and middle-income countries, the primary target population for a *Shigella* vaccine.

## INTRODUCTION

*Shigella* is a leading cause of moderate-to-severe diarrhea worldwide and the second cause of fatal diarrhea after rotavirus, with an estimated 148,000 deaths annually, including 93, 000 in children under 5, mostly in low-resource settings (1). Around 1.5-2 million new cases of shigellosis per year are also reported in high-income countries (1). Considering the global burden of shigellosis and the emerging antimicrobial resistance of *Shigella* (2, 3), a *Shigella* vaccine is of critical priority as recently defined by WHO (4).

The *Shigella* genus includes four serogroups or species: *S. dysenteriae* (15 serotypes), *S. boydii* (19 serotypes), *S. flexneri* (SF) (15 serotypes), and *S. sonnei* (Sson) with a single serotype, defined by carbohydrate composition of the repeating unit of the O-specific polysaccharide (O-SP) component of their lipopolysaccharide (LPS) (5). SF and Sson are prevalent worldwide and are responsible for approximately 90% of shigellosis cases (6, 7). SF, in particular serotypes 2a (SF2a) and 6 (SF6), are predominant in low- and middle-income countries (LMICs) (7) with sporadic episodes increasingly observed in high-income countries (1), while Sson is more prevalent in high-income and transitional countries (8–10).

Sero-epidemiological studies have shown that pre-existing serum IgG antibodies to Sson or SF2a LPS are strongly associated with resistance to homologous *Shigella* infection (11–13). This supported the notion that natural immunity conferred by pre-existing IgG antibodies was specific to the homologous bacteria, also underlining the immunodominant role of the O-SP, the most exposed surface antigen and the basis for serotyping. It was hypothesized that protection against *Shigella* infection might be conferred by vaccine-induced anti-O-SP serum IgG antibodies (14). On this basis, pioneering injectable conjugate vaccine candidates made of the detoxified LPS from SF2a, Sson and *S. dysenteriae* type 1, respectively, linked to carrier proteins using random conjugation chemistry, were developed in the early 90s by John B. Robbins and Rachel Schneerson at the National Institutes of Health (NIH) (15) paving the way to important developments in the field (16). Sson and SF2a detoxified LPS-rEPA (recombinant exoprotein A from *Pseudomonas aeruginosa*) conjugates – Sson-rEPA and SF2a-rEPA, respectively – induced high levels of serum IgGs against the homologous LPSs in phase I and II studies in healthy volunteers (15, 17). The vaccine efficacy (VE) of the Sson-rEPA conjugates was further demonstrated in field trials in Israel in young adults (74% VE after 1 dose) (18) and in children older than 3 years of age (71% VE after 2 doses) but not in the younger vaccine recipients (19). During the past decade, a new generation of glycoconjugates and other injectable LPS-based *Shigella* vaccines have been proposed which are currently in an advanced stage of clinical development to reach stronger immunogenicity and protection in infants (20–23). Going beyond the proof of concept in humans for monovalent vaccines, multivalent vaccine candidates combining various sets of O-SP antigens from Sson and SF serotypes 1b, 2a, 3a, and/or 6 were constructed (22, 24, 25) and others are being developed (26–28). The selection of SF serotypes to include in a multivalent *Shigell*a vaccine design is based on a combination of two complementary criteria, epidemiology data pointing to prevalent circulating strains on the one hand (7) and similarities or absence of similarities in SF O-SP protective epitopes on the other hand (29). The latter was supported by early studies of vaccine-induced cross-reactivity and cross-protection in guinea pigs (29, 30) and otherwise by molecular modeling (31) and more recent vaccine-induced cross-reactivity data in mice and rabbits (32).

Apart from SF6, all SF type-specific O-SPs share a common backbone repeating unit comprising four residues: three L-rhamnoses (Rha*p*) and a *N*-acetyl-D-glucosamine (Glc*p*NAc) (**Figure 1**). α-D-Glucopyranosyl, *O*-acetyl, and phosphoethanolamine groups occur under different patterns on this common tetrasaccharide, defining established SF antigenic type and group O-factors while also governing possible serotype cross-reactivity (33). Instead, the repeating unit of the SF6 O-SP backbone (**Figure 1**) consists of two Rha*p* residues, a galacturonic acid (Gal*p*A), and a *N*-acetyl-D-galactosamine (Gal*p*NAc) residue (34). As for several other SF O-SPs, including that from SF2a, the SF6 O-SP is 3/4-*O*-acetylated in a non-stoichiometric manner on a L-rhamnose residue. In fact, the occurrence of SF6 O-SPs featuring various levels *O*-acetylation led Perepelov el *et al.* to differentiate between SF6 and SF6a serotypes, corresponding to strains featuring low and high level of *O*-acetylation, respectively (35). This *O*-acetylation was found to be widespread among SF serotypes and to confer the host with a novel antigenic determinant provisionally named group factor 9 (36). Noriega et al. showed that a combined live attenuated SF2a and SF serotype 3a vaccine delivered intranasally to guinea pigs induced cross-reacting and cross-protecting secretory IgA and systemic IgG anti-LPS antibodies after intraoccular challenge with the homologous and heterologous SF serotypes (including SF1b, SF2b, SF4, and SF5) except SF6 (30). In recent years, the absence of cross-reactivity with SF6 O-SP of murine sera induced by SF2a O-SP-based vaccines was also reported in different studies (32, 37). However, detailed investigations showed that the advanced SF2a-rEPA conjugate vaccine developed at the NIH elicited antibodies cross-reacting with SF6 LPS and even suggested a trend of cross-protection (∼50%), against SF6 shigellosis in a clinical trial carried out in children in Israel (19, 37). The assumption was that the cross-reactivity/protection observed between the two serotypes is associated with the presence of the 3/4-*O*-acetyl-α-L-rhamnopyranosyl-(1→2)-α-L-rhamnopyranose segment (AB) that is part of the repeating units defining their O-SPs **(Figure 1)** (34, 37). A very recent study reported that immunization with the four-component GMMA-based vaccine altSonflex1-2-3, delivering *S. sonnei* and *S. flexneri* 1b, 2a, and 3a O-antigens, induced cross-reactive antibodies against SF6 O-SP in rabbits, minimal responses in rats, and no cross-reactivity in mice. In humans, functional cross-reactivity with SF6—defined as a ≥4-fold rise in serum bactericidal activity (SBA) titer—was observed in 0% and 22% of volunteers after one and two vaccine doses, respectively (38). These new findings have challenged the view on the lack of cross-reactivity between SF2a and SF6, also raising the question on the role of O-SP *O*-acetylation, in fact, non-stoichiometric *O*-acetylation, and questioned the relevance of animal models. On this background, we aimed to examine the potential cross-reactivity with SF6 LPS of antibodies induced by SF2a-TT15, the first synthetic glycan-based candidate vaccine developed against SF2a shigellosis (39) This novel monovalent sun-type glycoconjugate vaccine candidate features a chemically synthesized non-*O*-acetylated 15-mer oligosaccharide (OS) hapten linked through its reducing terminus via single-point attachment onto tetanus toxoid (TT) in an average OS:TT molar ratio of 17±5 (40, 41). The selected fine-tuned OS hapten corresponds to a three-basic-repeating-unit segment of the SF2a O-SP (31, 40–42). SF2a-TT15 was safe and well-tolerated and induced a strong immune response in a phase I study conducted in Israel in 2016-2017 (20). Despite the selection of a non-*O*-acetylated hapten in SF2a-TT15, sera induced in mice immunized with SF2a-TT15 recognized a large panel of SF2a circulating strains (39). We hypothesized that the strongly immunogenic SF2a vaccine candidate could induce cross-reacting antibodies to SF6 LPS in humans. To achieve the proof of concept, herein we examined the potential cross-reactivity with SF6 LPSs differing in their *O*-acetylation pattern, of IgG antibodies induced by SF2a-TT15 in young adult participants in the phase I clinical trial (20). The potential cross-reactivity was also evaluated against the LPS from *Escherichia coli* O147 (EC147), the O-SP of which is identical to a non-*O*-acetylated SF6 O-SP. We also assessed the extent to which cross-reacting anti-SF6 LPS IgG antibodies show functional capabilities, such as avidity and bactericidal activity against the heterologous LPSs, predicting potential cross-protection.

**Figure 1:**
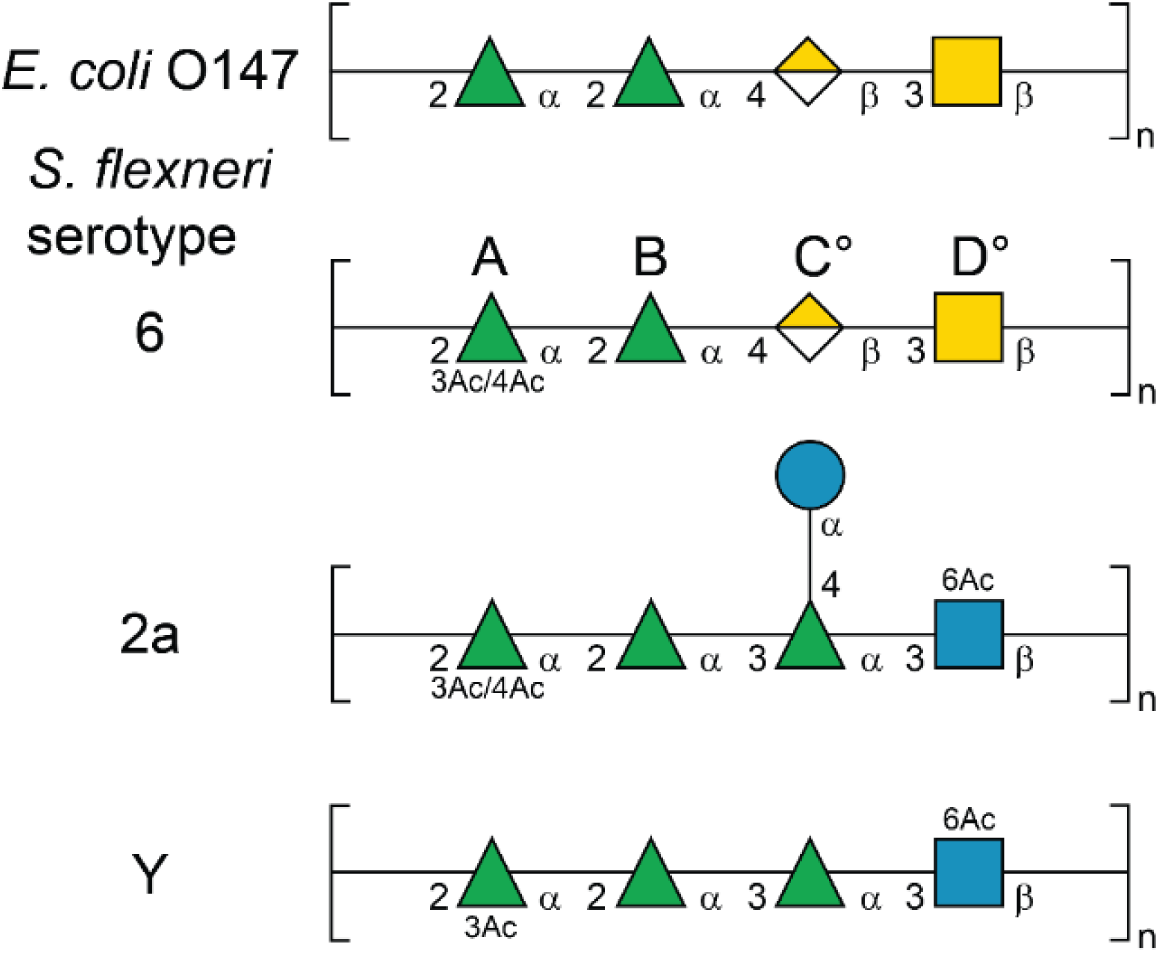
Schematic representation in SNFG format of the repeating unit of the O-Ag from *E. coli* O147(49) (43) SF2a and *S. flexneri* serotype Y (60). The sugar residues in SNFG format (61) were drawn by GlycanBuilder (62), and are denoted as follows: L-rhamnose is represented by a triangle filled with green color, D-galacturonic acid by a square half-filled with yellow color, *N*-acetyl-D-galactosamine by a square filled with yellow color, *N*-acetyl-D-glucosamine by a square filled with blue color, and D-glucose by a circle filled with blue color. Positions in sugar residues where *O*-acetylation may take place to various extent in different bacterial strains have been indicated. The specific *O*-acetylation content in each of the O-SP used in this study is indicated in Table 1. Ac: acetyl.

**Table 1:** *Shigella* strains used as a source of LPS, also showing the SF6 O-SP *O*-acetylation pattern and abbreviation.

| LPS description | % of *O*-acetylation at 3<sub>A</sub> / 4<sub>A</sub> | LPS abbreviation | Bacterial strain | Source |
| --- | --- | --- | --- | --- |
| *S. flexneri* 2a | 20% / 10%<sup>a</sup> | SF2a | 454 | Institut Pasteur, Centre National de Référence des Entérobactéries, France |
| *S. flexneri* 6<br>*O*-acetylated O-SP | 50% / 10%<sup>b</sup> | Ac-SF6 | 130316 | *Shigella* National Reference Laboratory, Ministry of Health, Israel |
| *S. flexneri* 6<br>low *O*-acetylated O-SP | 25% / 10%<sup>c</sup> | lowAc1- SF6 | Sc544 | Nils Carlin, Etvax, Sweden |
| *S. flexneri* 6<br>low-*O*-acetylated O-SP | 25% / 10%<sup>d</sup> | lowAc2-SF6 | MCDC 2924-71 | Institut Pasteur, Centre National de Référence des Entérobactéries, France |
| *S. flexneri* 6<br>non-*O*-acetylated O-SP | - / -<sup>b</sup> | noAc-SF6 | 75922 | *Shigella* National Reference Laboratory, Ministry of Health, Israel |
| *E. coli* O147<br>non-*O*-acetylated O-SP | - / -<sup>e</sup> | EC147 | *E. coli* O147<br>(serotype O147:H19: F4ac) | SSI Diagnostica, Denmark |
| *S. sonnei* O-SP | NA | Sson | NVGH1859 | GSK Vaccines Institute for Global Health, Italy |
| a. Ref 31 (Theillet et al), b. Ref 37 (Farzam et al), c. this work, d. Ref 43 (Chassagne et al), e. Ref 49 Hygge Blakeman K, et al.<br>NA = not applicable |  |  |  |  |

## METHODS

### Human serum samples

Serum samples from healthy adult volunteers participating in the SF2a-TT15 vaccine phase I study (20) were used for the examination of cross-reactivity of the SF2a-TT15-induced anti-SF2a LPS IgG with SF6 and EC147 LPSs. The design and results of the phase I study were described in detail elsewhere (20). Briefly, in a dose-escalating, single-blind, randomized, placebo-controlled study, 64 healthy adult volunteers (aged 18-45 years) were enrolled and assigned to receive either a low dose of the vaccine candidate, equivalent to 2 μg OS amount, or a high dose, equivalent to 10 μg OS amount, adjuvanted or not with aluminum hydroxide (alum), or matching placebo. Three single intramuscular injections were administered 28 days apart and participants were followed up for three months after the last injection. Cross-reactivity of the SF2a-TT15-induced anti-SF2a LPS IgG with SF6 and EC147 LPSs was examined twenty-eight days after receiving the second injection of the glycoconjugate.

### LPS and bacteria (see Table 1)

SF2a LPS was extracted from the SF2a strain 454 from Institut Pasteur, Centre National de Référence des Entérobactéries, Paris, France.

SF6 heterologous LPSs were extracted from strains 130316 (*O*-acetylated O-SP, Ac-SF6 LPS) and 75922 (non-*O*-acetylated O-SP, noAc-SF6 LPS) from the *Shigella* National Reference Laboratory, Ministry of Health, Jerusalem, Israel (37), strain Sc544 (low-*O*-acetylated O-SP, lowAc1-SF6 LPS), a gift from Nils Carlin, Etvax, Sweden and strain MCDC 2924-71 (low-*O*-acetylated O-SP, lowAc2-SF6 LPS), from Institut Pasteur, Centre National de Référence des Entérobactéries, Paris, France (43). Note that strains 130316 and 75922 are the two strains used in the pioneering study by Farzam S. et al. that first reported cross-reactivity of sera induced in human by a SF2a conjugate vaccine and SF6 LPS (37). A SF6-like LPS lacking the O-SP *O*-acetylation (35) was isolated from the EC147 reference strain (serotype O147:H19:F4ac, from SSI Diagnostica, Hillerød, Denmark). Sson LPS (a gift from GSK Vaccines Institute for Global Health, Siena, Italy) was isolated from strain NVGH1859 and used as a negative control LPS.

The LPSs of all *Shigella* and *E. coli* strains were extracted by the Westphal and Jann hot phenol-water extraction method, followed by dialysis and ultracentrifugation, as previously described (44).

### NMR spectroscopy of SF6 O-SP from strain Sc544

The LPS of SF6 strain Sc544 was treated with dilute acetic acid, and the resulting lipid-free polysaccharide was fractionated by size exclusion chromatography as previously described (43). NMR spectra were recorded in D_2_O at 298 K on Bruker AVANCE 500 MHz and AVANCE III 700 MHz spectrometers equipped with 5 mm Z-gradient CryoProbes. Chemical shifts are referenced relative to internal sodium 3-trimethylsilyl-(2,2,3,3-^2^H_4_)-propanoate (TSP, *δ*_H_ 0.00) and ^1^H NMR resonance assignments of isolated materials were carried out using standard NMR experiments for carbohydrates (45).

### Serum IgG antibodies to *Shigella* and EC147 LPSs

Serum IgG antibodies were measured by an ELISA in-house protocol (20). Briefly, 96 well microtiter plates (Corning Inc., New York, USA) were coated with 5 µg/mL of SF2a LPS, 0.5 µg/mL of Sson LPS, and 10 µg/mL from strains Sc544, MCDC 2924-71, 130316 and 75922 of SF6 LPSs with different degree of *O*-acetylation, and with 10 µg/mL of EC147 LPS in carbonate buffer for 1 h at 37 °C. These LPS concentrations gave the best discrimination between titers measured in pre- and post-infection or post-vaccination homologous paired sera.

Unbound sites were blocked for 1 h at 37 °C with 150 µL/well of blocking buffer containing 0.5% bovine serum albumin (BSA) (Merck Millipore Corp., Burlington, MA, USA) and 0.5% Casein (Sigma Aldrich, St. Louis, MO, USA). Wells were washed with phosphate buffered saline (PBS) (Biological Industries Ltd., Kibbutz Beit Haemek, Israel) containing 0.05% Tween-20 (Sigma Aldrich, St. Louis, MO, USA) and duplicates of tested samples were serially 2-fold diluted (initial dilution 1:200 for SF2a, SF6 and EC147 and initial dilution 1:100 for Sson) in blocking buffer, added to the wells and incubated overnight (ON) at room temperature (RT). Plates were washed and 100 µL/well of alkaline phosphatase (AP) conjugated anti-human IgG (Kirkegaard & Perry Laboratories Inc., Gaithersburg, MD, USA) diluted 1:5,000 in blocking buffer was added and incubated ON at RT. Wells were washed and 100 µL/well of phosphatase substrate, *para*-nitrophenyl phosphate (pNPP) one component (SouthernBiotech, Birmingham, AL, USA) was added and plates were incubated in the dark for 15 min at RT. The reaction was stopped by the addition of 50 µL/well of Stop solution (3M aqueous NaOH, Merck Millipore Corp., Burlington, MA, USA). Absorbance (A) was measured at 405 nm using an ELISA plate reader (Multiskan FC, Thermo Scientific, Waltham, MA, USA). Results were expressed in endpoint titers (the last serum dilution yielding an optical density (O.D.) of 0.2 or higher).

### Antibodies in lymphocyte supernatant (ALS)

Measurement of antibodies in ALS to SF2a LPS was done by ELISA on samples in 2-fold dilutions (20). Results were expressed in endpoint titers (the last supernatant dilution yielding a 0.2 or higher O.D). An ALS significant response was defined as a 4-fold or greater rise in titer in comparison with baseline when the second of the paired sample had an O.D. value of at least 0.2. IgG ALS cross-reactivity was measured after the first vaccine injection, when the homologous ALS levels were highest.

### Antibody avidity

Antibody avidity (functional affinity), defined as the overall binding strength of an antibody to an antigen was measured by competitive inhibition (46). Briefly, 96 well microtiter plates were coated with SF2a LPS or SF6 LPS, incubated for 1 h at 37 °C and blocked with 150 µL/well of blocking buffer (containing 0.5% BSA (Merck Millipore Corp., Burlington, MA, USA) and 0.5% Casein (Sigma Aldrich, St. Louis, MO, USA)) for another 1 h. Wells were washed with PBS (Biological Industries Ltd., Kibbutz Beit Haemek, Israel) containing 0.05% Tween-20 (Sigma Aldrich, St. Louis, MO, USA). Double dilutions of SF2a LPS or SF6 LPS, here as free antigen, in blocking buffer were added to the microtiter plate (initial concentration 5 µg/mL). The last well was covered only with the blocking buffer (control well). Sera at O.D. of approximately 1.0 at A405 were added to the wells containing the free antigen or blocking buffer, and plates were incubated ON. After washing the plates, 100 µL of a solution of AP-conjugated anti-human IgG (Kirkegaard & Perry Laboratories Inc., Gaithersburg, MD, USA) was added, followed by ON incubation. Plates were washed and 100 µL of pNPP one-component substrate (SouthernBiotech, Birmingham, AL, USA) was added and the plates were incubated in the dark for 15 min at RT. Plates were read at 405 nm until the O.D. of control wells reached 0.8 to 1.2 using an ELISA plate reader (Multiskan FC, Thermo Scientific, Waltham, MA, USA). Results are expressed as the log of the free antigen concentration that is necessary to cause 50% inhibition (*I*_50_).

### Serum Bactericidal Activity (SBA)

SBA was measured as previously reported (47, 48) with some modifications. Briefly, heat-inactivated serum samples were serially 2-fold diluted (initial dilution 1:50) in PBS (Biological Industries Ltd., Kibbutz Beit Haemek, Israel) in 96 U-shape microtiter plates (Greiner-bio-one GmbH, Kremsmünster, Austria) to a final volume of 20 µL/well. A bacterial suspension of SF2a, SF6s with different degrees of O-SP *O*-acetylation, or EC147 was grown in Tryptic Soy Broth (Becton, Dickinson and Company, Franklin Lakes, NJ, USA) supplemented with 0.4% Yeast Extract (HiMedia Laboratories Pvt. Ltd., Mumbai, India) at 37 °C in a shaker incubator to its logarithmic phase (O.D 0.3-0.4) at 600 nm depending on the bacterial suspension; ∼7x10^7^ cfu/mL). The suspension was diluted 1:10,000, and 40 µL/well was added to the microtiter plates. Plates were placed in a shaker incubator at 37 °C for 15 min followed by the addition of 40 µL/well (40% complement) of Baby Rabbit Complement (BRC) (Cedarlane Corporation, Burlington, ON, Canada), the maximum BRC concentration not causing non-specific bacterial killing and 1-2 h incubation at 37 °C without shaking. Negative-control wells containing only bacteria and BRC (without serum) were included in each assay. After incubation, 20 µL of the samples from each well were diluted in 380 µL of PBS followed by plating six times 20 µL drops on Tryptic Soy Agar-Congo Red plates (Hy-Laboratories LTD, Rehovot, Israel). The number of colonies was counted and the percentage of bacteria survival compared to control wells (containing bacteria and BRC without serum) was calculated. A serum yielding at least 50% reduction in the number of colonies compared to control wells was defined as having bactericidal activity. The last serum dilution yielding bactericidal activity was defined as the end point titer.

### Statistical analysis

Geometric mean titer (GMT) and GMT ratio (GMTR) e.g., titer on day 56 vs baseline, and 95% confidence intervals (CIs) were calculated for the immunological parameters measured. The percentages of responders to homologous and heterologous LPSs were determined using a priori-defined cutoff values for each immunological parameter. Student’s t-Test was used to examine differences in mean and GMT of binding and functional antibodies against homologous (SF2a) and heterologous (SF6, Sson and EC147) LPSs. Fisher’s exact test was employed to analyze differences in the percentage of responders to homologous and heterologous LPSs. All statistical tests were interpreted in a two-tailed fashion using a significance level (α) = 0.05. Data were analyzed using the SPSS version 24 (Armok, New York, USA).

## RESULTS

We examined the IgG response of recipients of the 2 µg OS and 10 µg OS doses of the SF2a-TT15 vaccine candidate or placebo to the homologous SF2a LPS, heterologous SF6 and EC147 LPS preparations, and a control Sson LPS. Special focus was on the role of the O-SP *O*-acetylation (O-SPAc) in inducing cross-reactivity between SF2a and SF6 LPSs. To this end, we studied the binding of SF2a-TT15 induced sera to LPSs isolated from four SF6 strains (**Table 1**), which differ by their O-SP *O*-acetylation pattern, and to an EC147 LPS, whose O-SP is identical to the SF6 O-SP backbone, albeit lacking the *O*-acetyl substitutions (49).

### Analysis of the O-SP component of SF6 LPSs

The LPS from SF6 strain Sc544 was first analyzed by ^1^H NMR spectroscopy that showed, inter alia, resonances at ∼2.05 ppm anticipated to originate from the methyl group of the *N*-acetyl group of the D-Gal*p*NAc residue in the repeating unit and, in addition, peaks at ∼2.2 ppm indicating the presence of *O*-acetyl groups as substituents on the O-SP (35).

From the treatment of the LPS with dilute acetic acid, followed by size exclusion chromatography, a polymeric fraction and an oligomeric fraction were selected for further analysis by NMR spectroscopy. The ^1^H NMR spectrum of the polymeric material was similar to that of the O-SP from SF6 strain MCDC 2924-71 (lowAc2-SF6), and the degree of *O*-acetylation was determined by integration of resonances at *δ*_H_ 2.21 ppm from 3_A_-H and *δ*_H_ 2.16 ppm from 4_A_-H vs. *N*-acetyl groups of residue B at *δ*_H_ 2.08 ppm due to *O*-acetylation at 3_A_, at *δ*_H_ 2.04 ppm devoid of *O*-acetyl groups in the repeating unit and at *δ*_H_ 2.03 ppm due to *O*-acetylation at 4_A_, which revealed the former to be *O*-acetylated to 1/4 and the latter to 1/10 of the repeating units of the polymeric chain (**Figure 2a**), closely similar to the O-SP from SF6 strain MCDC 2924-71 (43). The *O*-acetylation pattern of the O-SP composing the polymeric material was confirmed by ^1^H,^1^H-TOCSY NMR experiments with correlations from the H6 protons of methyl groups of rhamnose residues A and B (**Figure 2c**). The oligomeric material also showed that the rhamnobiose segment was *O*-acetylated at positions 3 and 4 of residue A, but in addition and to a significant extent, revealed *O*-acetylation at position 2 (**Figure 2b**) of the terminal sugar residue of the O-SP biological repeating unit [_Ac_ABC°D°] (**Figure 1**), for which the resonance assignment was further aided by the ^1^H,^1^H-TOCSY NMR spectra (**Figure 2d**) employing an array of mixing times with a duration of 10 – 120 ms. Besides a single repeating unit, the oligomeric material also contains short O-SP chains with an additional repeating unit evident from the similarity of e.g. *O*-acetylation at 3_A_ (cf. horizontal line between **panels c and d in Figure 2**). That the ^1^H,^1^H-TOCSY trace revealing a cross-peak at, *inter alia*, *δ*_H_ 5.23 ppm corresponds to *O*-acetylation at 2_A_ is supported by a ^1^H NMR chemical shift prediction using the CASPER program (50, 51) which using a model for the _Ac_AB terminal part of the repeating unit, α-L-Rha*p*_2Ac_-(1→2)-α-L-Rha*p*-OMe, showed for its terminal residue *δ*_H2_ 5.24 ppm, in excellent agreement with the experimentally observed chemical shift. One may note that the ^1^H NMR chemical shifts of the spin systems for *O*-acetylation at 4_A_ is similar in the long chain O-SPs and the short ones, but not identical, as evident from **panels c and d of Figure 2**, e.g. *δ*_H_ 1.15 ppm and 4.82 ppm in the former but *δ*_H_ 1.16 ppm and 4.88 ppm in the latter. Furthermore, the ^1^H NMR chemical shifts of the terminal rhamnose residue A are in good agreement with those reported for a one repeating unit-core oligosaccharide from a semi-rough SF6 LPS (52).

**Figure 2.**
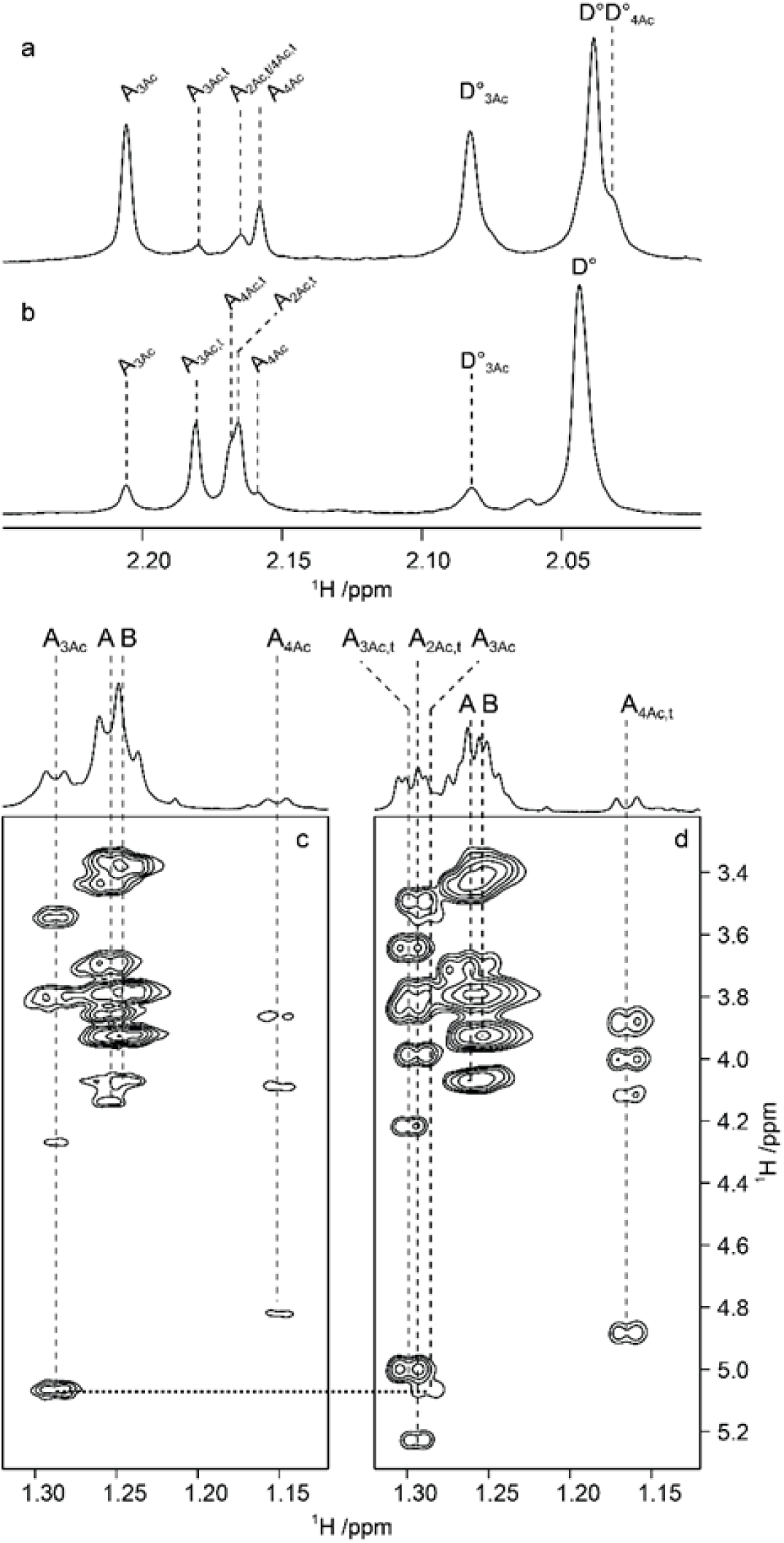
Selected regions of ^1^H NMR spectra of (a) the O-SP and (b) the oligomeric material from SF6 strain Sc544, showing resonances from *O*- and *N*-acetyl groups; ^1^H,^1^H-TOCSY (τ_mix_ 120 ms) NMR spectra of (c) the polysaccharide material and (d) the oligomeric material showing spin systems originating from the H6 protons in the methyl groups of the rhamnosyl residues. Annotations are given with respect to the *O*-acetylation patterns in the polymeric and oligomeric materials, respectively. The horizontal dotted line between the two panels highlights the H6/H3 cross-peak from residue A, a sugar that is *O*-acetylated in position 3.

### SF2a-TT15 induces serum IgG antibodies cross-reacting with heterologous SF6 LPSs

There was a 4-fold or greater rise in anti-SF2a LPS serum IgG levels between pre-vaccination (day 0) and 28 days after the second injection (day 56 after the first injection) in 83% and 96% of the volunteers, who had received the 2 and 10 µg OS doses of SF2a-TT15, respectively (**Table 2**). There was a 4-fold or greater rise in IgG levels against heterologous SF6 LPSs elicited by the SF2a-TT15 vaccine candidate, with a higher percentage of responders in the group of volunteers who were vaccinated with the 10 µg OS dose compared with those who received the 2 µg OS vaccine dose (**Table 2**). Cross-reactivity was consistently higher with the lowAc1-SF6, lowAc2-SF6 and EC147 LPS preparations (highest with the lowAc1-SF6 LPS) as compared with that measured against the Ac-SF6 LPS (**Table 2**).

**Table 2:** Phase I study volunteers with anti-SF2a LPS, SF6 LPSs, EC147 LPS and Sson LPS serum IgG titers showing ≥ 4-fold rise following two injections of SF2a-TT15 or placebo.

| Cohort | Vaccine recipients |  |  |  |  |  |  | Placebo recipients |  |  |  |  |  |  |
| --- | --- | --- | --- | --- | --- | --- | --- | --- | --- | --- | --- | --- | --- | --- |
|  | SF2a LPS | lowAc1-SF6 LPS<br>(Sc544 strain) | lowAc2-SF6 LPS<br>(MCDC 2924-71 strain) | Ac-SF6 LPS<br>(130316 strain) | NoAc-SF6 LPS<br>(75922 strain) | EC147 LPS<br>(O147: H19:F4ac strain) | Sson LPS | SF2a LPS | lowAc1-SF6 LPS<br>(Sc544 strain) | lowAc2-SF6 LPS<br>(MCDC 2924-71 strain) | Ac-SF6 LPS<br>(130316 strain) | NoAc-SF6 LPS<br>(75922 strain) | EC147 LPS<br>(O147: H19:F4ac strain) | Sson LPS |
|  | N (%) | N (%) | N (%) | N (%) | N (%) | N (%) | N (%) | N (%) | N (%) | N (%) | N (%) | N (%) | N (%) | N (%) |
| **Cohort 1** | 19/23<br>(83) | 5/23<br>(22) | 3/23<br>(13) | 1/23<br>(4) | 3/23<br>(13) | 3/9<br>(33) | 0/23<br>(0) | 0/8<br>(0) | 0/8<br>(0) | 0/8<br>(0) | 0/6<br>(0) | 0/8<br>(0) | ND | 0/8 (0) |
| **Cohort 2** | 23/24<br>(96) | 12/24<br>(50) | 7/24<br>(29) | 5/24<br>(21) | 12/24<br>(50) | 8/24<br>(33) | 0/24<br>(0) | 0/8<br>(0) | 0/8<br>(0) | 0/8<br>(0) | 0/8<br>(0) | 0/8<br>(0) | ND | 0/8 (0) |

N: number of sera tested
vaccine recipients: cohort 1: 2 µg OS dose of SF2a-TT15; cohort 2: 10 µg OS dose of SF2a-TT15

Of the vaccinees receiving the 2 µg OS dose, 4% to 33% had a 4-fold or greater rise in IgG against SF6 LPSs and EC147 LPS. Of the vaccinees receiving the 10 µg OS dose, 21-50% had a 4-fold or greater rise in IgG against SF6 LPSs and EC147 LPS (**Table 2**). None of the volunteers receiving the low or high dose of SF2a-TT15 had a 4-fold or higher rise in IgG against Sson LPS 28 days after the second injection (**Table 2**). Likewise, none of the 16 placebo recipients showed any rise in IgG titer against SF2a LPS or SF6 LPSs or Sson LPS documenting no natural exposure to *Shigella* during the study period.

The magnitude (GMT) of the IgG response to homologous and heterologous LPSs in the volunteers receiving the SF2a-TT15 vaccine candidate is shown in **Table S1**. Twenty-eight days after receiving the second injection of the glycoconjugate, the rises of GMTs against homologous SF2a LPS and all four heterologous SF6 LPSs were more pronounced after vaccination with the 10 µg OS vaccine dose as compared to the 2 µg OS vaccine dose (**Table S1**). The GMTR of IgG antibodies between post-10 µg OS vaccination versus pre-vaccination, were 26.1, 3.8, 3.5, 2.3, 1.7 and 2.9 against SF2a, lowAc1-SF6, noAc-SF6, lowAc2-SF6, Ac-SF6 and EC147 LPSs, respectively (**Table S1**). The GMTR of IgG antibodies between post-10 µg OS vaccination versus pre-vaccination, in volunteers with a 4-fold or greater rise in IgG, were 29.2, 8.0, 8.0, 6.6, 4.6 and 7.3 against SF2a, lowAc1-SF6, noAc-SF6, lowAc2-SF6, Ac-SF6 and EC147 LPSs, respectively (**Table 3**). Consistent with the results presented in **Table 2**, the highest immune response by GMTR against SF6 and SF6-like LPSs was observed for the lowAc1-SF6 LPS. Moreover, the immune response estimated by GMTR for the Ac-SF6 LPS was non-existent or lower than that to any of the lowAc-SF6 and noAc-SF6 LPSs, as illustrated for the MCDC 2924-71 LPS (lowAc2-SF6 LPS), the 75922 LPS (noAc-SF6 LPS) and theEC147 LPS featuring a non-*O*-acetylated SF6 O-SP (**Table S1**). As expected, no rise in serum IgG titer was detected against Sson LPS following SF2a-TT15 vaccination.

**Table 3:**
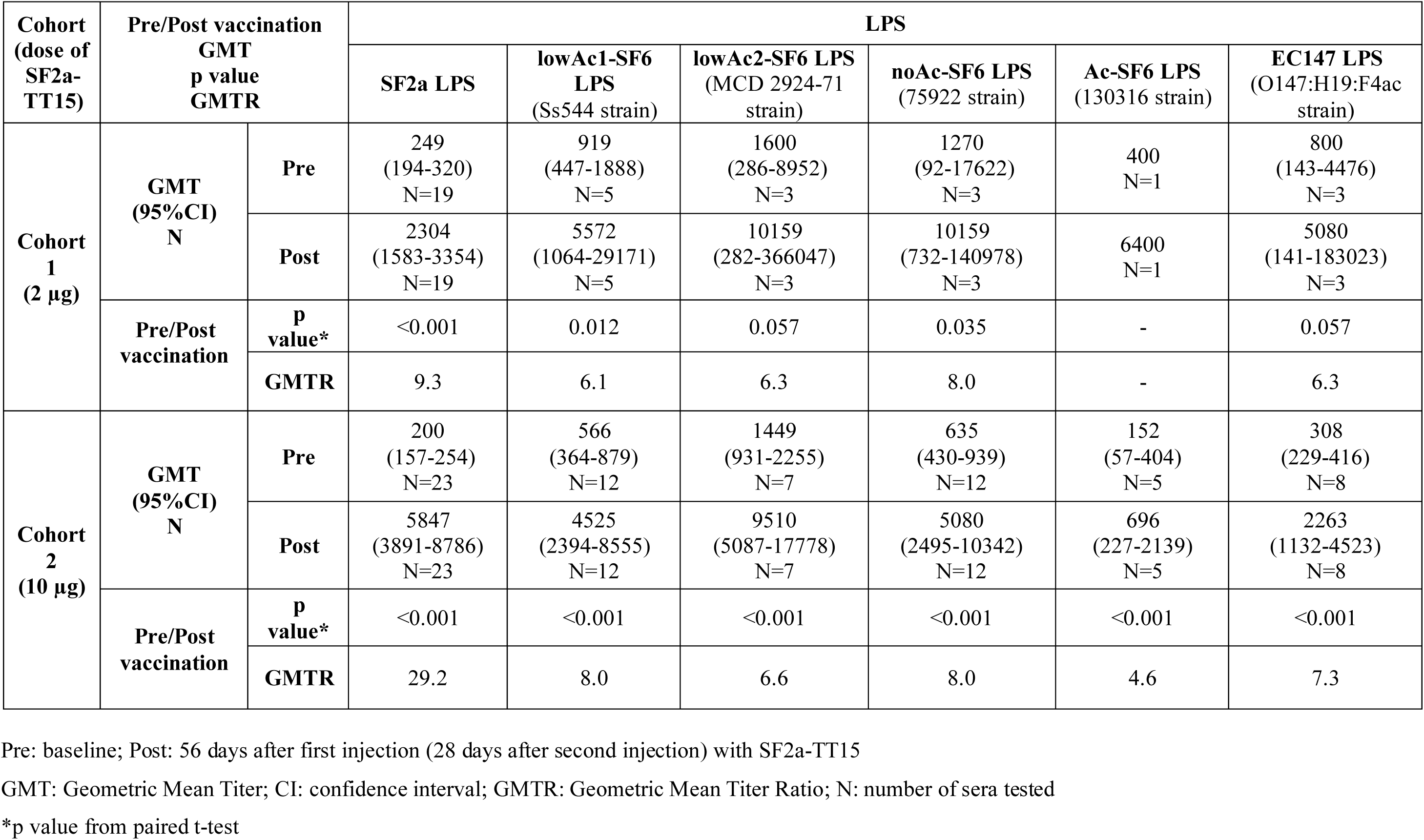
Serum IgG levels against SF2a LPS, SF6 LPSs, EC147 LPS and Sson LPS following two injections of SF2a-TT15 in vaccinees with a ≥ 4-fold rise in titer between baseline and 28 days after the second injection.

### SF2a-TT15 induces IgG ALS cross-reacting with heterologous SF6 LPSs

IgG antibodies to the SF2a LPS in lymphocyte supernatant (ALS) induced by the first injection of the SF2a-TT15 vaccine candidate cross-reacted with the lowAc1-SF6 LPS (Sc544 strain) when assessed in a subsample of volunteers. Two out of 6 volunteers (33.3%) immunized with the 2 µg OS dose of SF2a-TT15 and 11 of 18 volunteers (61.1%) immunized with the 10 µg OS dose had a ≥ 4-fold rise in ALS to lowAc1-SF6 LPS with 4.4- and 13.8-fold rise in the corresponding GMTs between pre-vaccination and day 7 after the first SF2a-TT15 injection, respectively. No significant rise in IgG antibodies to SF2a LPS and lowAc1-SF6 LPS in ALS was detected among the six tested placebo recipients (**Table 4**).

**Table 4:** Levels of anti-SF2a LPS and anti-SF6 Sc544 LPS (lowAc1-SF6 LPS) antibodies in lymphocyte supernatant (ALS) and percent of ALS responders* 7 days following the administration of the first dose of SF2a-TT15 vaccine candidate or placebo.

| Cohort | Pre/Post<br>vaccination<br>GMT<br>p value<br>GMTR<br>N (%) | SF2a-TT15 recipients |  | Placebo recipients |  |
| --- | --- | --- | --- | --- | --- |
|  |  | SF2a LPS | lowAc1-SF6 LPS<br>(Sc544 strain) | SF2a LPS | lowAc1-SF6 LPS<br>(Sc544 strain) |
|  |  | GMT (95%CI)<br>N | GMT (95%CI)<br>N | GMT (95%CI)<br>N | GMT (95%CI)<br>N |
| Cohort 1 | Pre | 0.66 (0.66-0.66)<br>N=24 | 0.66 (0.66-0.66)<br>N=6 | 0.66 (0.66-0.66)<br>N=8 | ND |
|  | Post | 4.34 (2.06-9.14)<br>N=24 | 2.87 (0.59-13.95)<br>N=6 | 0.66 (0.66-0.66)<br>N=8 | ND |
|  | p value # | < 0.001 | 0.062 | 1.0 | ND |
|  | GMTR | 6.6 | 4.4 | 1.0 | ND |
|  | N (%) with<br>≥4-fold rise* | 12 (50.0%) | 2 (33.3%) | 0 (0%) | ND |
| Cohort 2 | Pre | 0.66 (0.66-0.66)<br>N=24 | 0.66 (0.66-0.66)<br>N=18 | 0.66 (0.66-0.66)<br>N=8 | 0.66 (0.66-0.66)<br>N=6 |
|  | Post | 34.15 (14.38-81.08)<br>N=24 | 9.12 (3.67-22.68)<br>N=18 | 0.66 (0.66-0.66)<br>N=8 | 0.66 (0.66-0.66)<br>N=6 |
|  | p value # | < 0.001 | < 0.001 | - | - |
|  | GMTR | 51.7 | 13.8 | 1.0 | 1.0 |
|  | N (%) with<br>≥4-fold rise* | 23 (95.8%) | 11 (61.1%) | 0 (0%) | 0 (0%) |

GMT: Geometric Mean Titer; CI: confidence interval; GMTR: Geometric Mean Titer Ratio; Pre: baseline; Post: 7 days after first injection of SF2a-TT15
\* ≥4-fold rise in titer between pre-vaccination and 7 days after first dose

### Functional capabilities of SF2a-TT15-induced serum IgGs towards SF2a and SF6 strains

**Table 5** displays the killing capabilities of sera of volunteers obtained 28 days after the second injection of the 2 µg and 10 µg OS doses of the SF2a-TT15 vaccine candidate against the homologous SF2a 2457T strain and various heterologous SF6 and EC147 strains. Killing was evaluated in vaccinees with a 4-fold or greater rise in serum IgG titers to the corresponding LPS. A 4-fold or greater rise in SBA titer was found against SF2a 2457T strain in 68% (13/19) and 91% (21/23) of the volunteers receiving the 2 µg and 10 µg OS doses of SF2a-TT15, respectively. Under the same SBA assay conditions, 14% to 58% of sera of SF2a-TT15 vaccinees with a 4-fold or greater rise in serum IgG titers against the heterologous SF6 or EC147 LPSs also yielded 4-fold or greater rises in SBA titer when tested against the corresponding strains (**Table 5**).

**Table 5:**
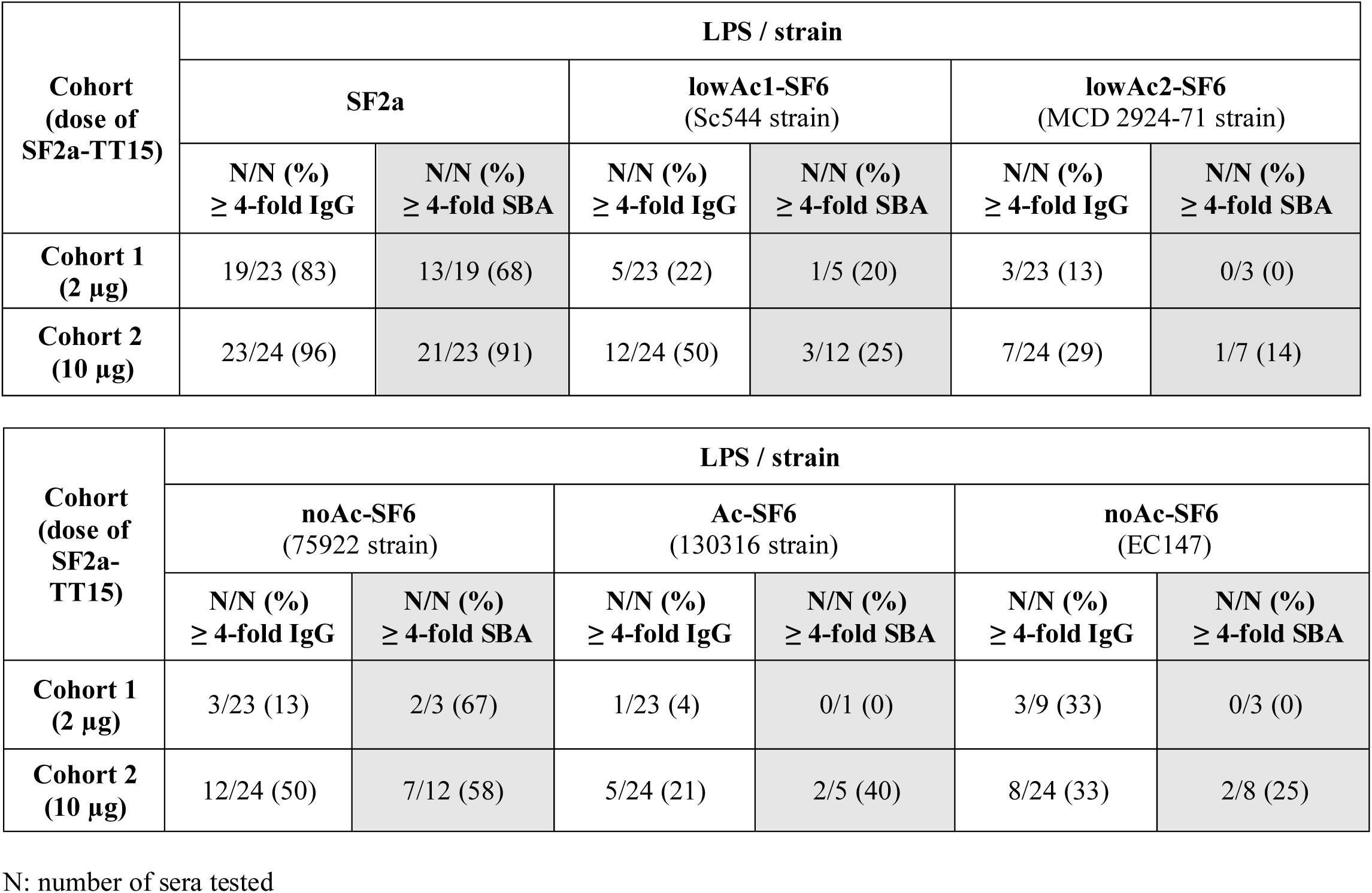
Percent of responders with a ≥ 4-fold rise in serum IgG titers to SF2a LPS, SF6 LPSs and EC17 LPS in vaccinees and for each one, % of responders with a ≥ 4-fold rise in SBA between baseline and 28 days after the second injection of SF2a-TT15.

| Cohort<br>(dose of<br>SF2a-TT15) | LPS / strain |  |  |  |  |  |
| --- | --- | --- | --- | --- | --- | --- |
|  | SF2a |  | lowAc1-SF6<br>(Sc544 strain) |  | lowAc2-SF6<br>(MCD 2924-71 strain) |  |
| | N/N (%)<br>$\geq 4$-fold IgG | N/N (%)<br>$\geq 4$-fold SBA | N/N (%)<br>$\geq 4$-fold IgG | N/N (%)<br>$\geq 4$-fold SBA | N/N (%)<br>$\geq 4$-fold IgG | N/N (%)<br>$\geq 4$-fold SBA |
| **Cohort 1**<br>(2 μg) | 19/23 (83) | 13/19 (68) | 5/23 (22) | 1/5 (20) | 3/23 (13) | 0/3 (0) |
| **Cohort 2**<br>(10 μg) | 23/24 (96) | 21/23 (91) | 12/24 (50) | 3/12 (25) | 7/24 (29) | 1/7 (14) |

| Cohort<br>(dose of<br>SF2a-TT15) | LPS / strain |  |  |  |  |  |
| --- | --- | --- | --- | --- | --- | --- |
|  | noAc-SF6<br>(75922 strain) |  | Ac-SF6<br>(130316 strain) |  | noAc-SF6<br>(EC147) |  |
| | N/N (%)<br>$\geq 4$-fold IgG | N/N (%)<br>$\geq 4$-fold SBA | N/N (%)<br>$\geq 4$-fold IgG | N/N (%)<br>$\geq 4$-fold SBA | N/N (%)<br>$\geq 4$-fold IgG | N/N (%)<br>$\geq 4$-fold SBA |
| **Cohort 1**<br>(2 μg) | 3/23 (13) | 2/3 (67) | 1/23 (4) | 0/1 (0) | 3/9 (33) | 0/3 (0) |
| **Cohort 2**<br>(10 μg) | 12/24 (50) | 7/12 (58) | 5/24 (21) | 2/5 (40) | 8/24 (33) | 2/8 (25) |

N: number of sera tested

We measured the avidity of serum IgG antibodies to SF2a and to the low Ac1-SF6 LPS (Sc544 strain), which showed the highest cross-reactivity of IgG anti-LPS antibodies. This was done in volunteers after receiving two injections of SF2a-TT15 at 10 µg OS and in placebo recipients. For both the homologous (SF2a) and the heterologous antigen (SF6), a significant increase in avidity in sera of vaccinees receiving the 10 µg OS dose of SF2a-TT15 was found, 28 days after the second injection, in comparison to pre-immune sera (**Table 6**). No difference in avidity was found between baseline and post-immunization sera of placebo recipients, as evaluated against both SF2a and low Ac1-SF6 LPSs.

**Table 6:** Avidity (expressed in Mean *I*_50_ (CI 95%)) of serum IgGs to SF2a LPS and SF6 lowAc1 LPS following one and two injections of the 10 µg OS dose of SF2a-TT15 or placebo.

| Pre/Post<br>vaccination<br>p value | 10 µg OS dose SF2a-TT15 recipients |  |  | Placebo recipients |  |  |
| --- | --- | --- | --- | --- | --- | --- |
| | N | SF2a LPS<br>Mean $I_{50}$ (95% CI) | lowAc1-SF6 LPS Sc544<br>Mean $I_{50}$ (95% CI) | N | SF2a LPS<br>Mean $I_{50}$ (95% CI) | lowAc1-SF6 LPS Sc544<br>Mean $I_{50}$ (95% CI) |
| **Pre** | 12 | 1.81 (1.43-2.19) | 2.15 (1.65-2.65) | 4 | 2.34 (1.11-3.57) | 2.15 (0.59-3.71) |
| **Post 1** | 12 | 1.34 (1.2-1.49) | 1.52 (1.15-1.88) | 4 | 2.37 (1.19-3.55) | 2.17 (0.63-3.71) |
| **Post 2** | 12 | 1.29 (1.18-1.41) | 1.61 (1.23-1.98) | 4 | 2.4 (1.27-3.53) | 2.16 (0.59-3.72) |
| **p value**<br>**(Pre vs Post 1)\*** |  | 0.030 | 0.008 |  | 0.192 | 0.709 |
| **p value**<br>**(Pre vs Post 2)\*** |  | 0.019 | 0.016 |  | 0.234 | 0.876 |

\*Paired t test between Pre and Post-vaccination
N: number of samples
CI: confidence interval
Pre: baseline; Post 1: 28 days after the first injection; Post 2: 28 days after the second injection (56 days after the first injection) with SF2a-TT15

## DISCUSSION

This study investigated the possible cross-reactivity between heterologous SF6 LPSs and antibodies induced in young adult volunteers by SF2a-TT15, a novel conjugate vaccine candidate against SF2a shigellosis and the first one made of chemically synthesized haptens. Cross-reactivity of binding and functional antibodies was assessed after two vaccine injections, as a primary 2-dose vaccination regimen is the recommended regimen in a *Shigella* vaccine preferred product characteristics (53). It is also the regimen evaluated in phase IIa and IIb studies of the SF2a-TT15 vaccine candidate (ClinicalStudies.gov NCT04602975 and NCT04078022), though on different intervals between the first and second doses.

### Vaccine design and implications for cross-reactivity

SF2a-TT15 is unique among several SF2a LPS-based vaccine candidates that have been proposed owing to the well-defined synthetic 15mer OS, a non-natural albeit functional SF2a O-SP surrogate, designed for use as hapten (39). In contrast to polysaccharide conjugate vaccines derived from extracted LPS, the LPS core is absent from SF2a-TT15. Therefore, the immune response induced against SF2a is directed at the O-SP only, and the same is assumed for any observed cross-reactivity with heterologous LPSs. It is also noteworthy that following the demonstration by us (54) and others (35) that the naturally occurring O-SP non-stoichiometric *O*-acetylation was not critical for antigenicity, the selected glycan hapten component in SF2a-TT15 corresponds to a three non-*O*-acetylated repeating unit segment of the SF2a O-SP (55). Subsequent reports by us (39) and others (34, 56) demonstrating no major impact of *O*-acetyl decorations on the immune response, supported the original SF2a-TT15 design.

### Evidence of cross-reactivity in humans

We report here that the SF2a-TT15-induced antibodies in humans did cross-react with SF6 LPSs. The observed cross-reactivity correlated with the magnitude of the IgG antibody response to the homologous LPS, indicating that robust homologous immunity is a key determinant of heterologous recognition. Participants in the phase I clinical trial of the SF2a-TT15 vaccine candidate were adults with low baseline titers of anti-SF2a LPS serum IgG antibodies recruited from the Tel Aviv area and surroundings, where the risk of exposure to SF2a and SF6 is very low. The herein obtained data, showing no increase in IgG titer against the SF2a LPS or the SF6 LPS compared with baseline in placebo recipients, documented the absence of natural exposure to either of these two SF serotypes or to other cross-reacting antigens in the study population during the follow-up period. It follows that the SF2a-TT15 vaccine candidate induced both homologous and cross-reactive immune responses, as disclosed in the present analysis.

Importantly, the same cross-reactivity was detected in IgG antibodies measured in ALS at day 7 after vaccination. Since ALS reflects antibodies secreted by recently activated circulating plasmablasts, this finding provides direct B-cell level evidence that vaccination induces cross-reactive B-cell responses. The higher proportion of ≥4-fold ALS responders and the greater GMT rise in the 10 µg OS group compared with the 2 µg group demonstrate a clear dose response relationship. Together with the absence of such responses in placebo recipients, these results confirm that the cross-reactivity is vaccine-driven.

### Functional properties of cross-reactive antibodies

The cross-reacting antibodies against SF6 LPSs induced by SF2a-TT15 were partially functional, demonstrating bactericidal activity and increased avidity for the heterologous LPSs. Among SF2a-TT15 vaccinees, 14%–58% of sera with a ≥4-fold increase in IgG titers to heterologous SF6 or EC147 LPS also showed a ≥4-fold rise in SBA titers against the corresponding strains. This could be the result of a greater susceptibility of SF2a strains to the bactericidal activity of human serum compared to SF6 strains, as previously reported (57) and also observed by us following natural infection with the two serotypes (data not shown). Differences in epitope specificity between binding and bactericidal antibodies could be an additional explanation. For both the homologous (SF2a) and the heterologous antigen (SF6), a significant increase in avidity in sera of vaccinees receiving the 10 µg OS dose of SF2a-TT15 was found, 28 days after the second injection, in comparison to pre-immune sera while no increase in avidity was observed among placebo recipients.

### Comparison with other vaccine platforms

To the best of our knowledge, this is the second report on the cross-reactivity between IgG antibodies to SF2a LPS and SF6 LPSs in humans. The rate observed in this study is higher than the approximate 20% cross-reactivity with Ac-SF6 LPS (130316 strain) previously shown in human recipients of the SF2a-rEPA conjugate at a dose containing 25 µg O-SP (37). Interestingly, similar cross-reactivity (21%) with the same Ac-SF6 LPS (130316 strain) was observed in this study among recipients of the SF2a-TT15 vaccine candidate despite a different glycoconjugate composition. An additional very recent study reported that post-immunization sera from European adults immunized with the 4-component GMMA-based vaccine altSonflex1-2-3, delivering Sson, SF1b, SF2a and SF3a O-antigens (GMMA also containing the core and not only the O-SP of the corresponding LPS) induced cross-reactive functional antibodies with SF6 (4-fold or greater rise in SBA titer) in 0 and 22% of volunteers’ sera after one or two injections, respectively (38). These human findings contrast with earlier mouse and guinea pig data reporting the absence of cross-reactivity between SF2a and SF6 but align with more recent evidence of heterologous bactericidal activity observed in rabbits immunized with GMMA (32, 38). Together, these results highlight the importance of species selection and vaccine platform when evaluating cross-reactivity.

The functional parameters, such as bactericidal activity and avidity of the cross-reacting antibodies, demonstrated in some of the volunteers suggest that these antibodies could confer potential cross-protection when induced by vaccination, especially with a vaccine candidate comprising a strongly immunogenic SF2a component.

### Influence of *O*-acetylation

Cross-reactivity appeared to be dependent on the O-SP *O*-acetylation level. IgG cross-reacting antibodies induced by the SF2a-TT15 vaccine candidate against heterologous lowAc-SF6 LPSs raised by a ≥ 4-fold compared to baseline titers in more than 50% of the volunteers that had received the higher dose of the vaccine, either adjuvanted or not. In contrast to previous assumptions (31), our findings suggest that the cross-reactivity between SF2a and SF6 O-SPs observed in SF2a-TT15 recipients is not necessarily related to the non-stoichiometric 3/4-*O*-acetylation of rhamnose A naturally present in a random fashion along the basal O-SPs (**Figure 1**). Indeed, herein cross-reactivity was observed with all the SF6 O-SPs that were tested. Yet, the strongest cross-reactivity was directed at the lowAc-SF6 and noAc-SF6 LPSs compared to the Ac-SF6 LPS. This is not unexpected as SF2a-TT15 features by design a non-*O*-acetylated synthetic SF2a O-SP segment as native LPS surrogate. Along the same vein, this original vaccine candidate can induce antibodies against a wide diversity of SF2a circulating strains, which are expected to differ in their *O*-acetylation patterns (39). In agreement with our findings, recent conformational analysis by means of molecular dynamics simulations of SF6 six repeating unit O-SP segments, lacking *O*-acetylation, 100% 3_A_-*O*-acetylated or 100% 4_A_-*O*-acetylated, respectively, showed that *O*-acetylation has little effect on the SF6 O-SP conformation and hence may not be essential for antigenicity and therefore cross-reactivity (34). Otherwise and as already observed in mice for polysaccharide-CRM_197_ conjugates made of the natural SF6 polysaccharide displaying 48% and 18% *O*-acetylation at positions 3 and 4 of rhamnose A, respectively, or of its de-*O*-acetylated counterpart (58), immunogenicity data in mice comparing GMMA (Generalized Modules for Membrane Antigens) from wild-type SF6 displaying a fully *O*-acetylated polysaccharide antigen (73% at O-3_A_ and 17% at O-4_A_) and a mutant thereof displaying a noAc-SF6 polysaccharide antigen, confirmed that *O*-acetylation does not play a major role on the ability of an O-SP-based SF6 vaccine at inducing a functional homologous antibody response against both Ac-SF6 and noAc-SF6 bacteria (34). We are not ruling out the possible role of the *O*-acetyl groups in the cross-reactivity induced by a naturally *O*-acetylated O-SP conjugate vaccine, but we raise the likelihood that other epitopes displayed in a noAc-SF2a O-SP-based vaccine candidate might lead to a significant immune response to the heterologous SF6 LPSs when a strong immunity is achieved. Based on our results, we postulate that the strong homologous response induced by SF2a-TT15 in humans determined the cross-reactivity with the heterologous SF6 LPSs. In accordance with this hypothesis, albeit going beyond an established dogma, are findings from the extensive analysis of cross-reactivity of antisera raised in mice against GMMA from one SF subtype to heterologous SF serotypes. In contrast to expectations, most observed cross-reactivity could not be assigned to the well-known O-SP modifications that define SF types and subtypes, suggesting that cross-reactive epitopes in humans have yet to be defined on a molecular basis (32).

There are several possible explanations for the diverging findings on the absence of cross-reactivity and cross-protection between anti-SF2a LPS IgG antibodies and SF6 bacteria in the guinea pig model (30) and the observed cross-reactivity reported in humans following immunization with the SF2a *O*-acetylated O-SP-based SF2a-rEPA conjugate (37) and the non-*O*-acetylated synthetic OS-based SF2a-TT15 conjugates as demonstrated in the present study. Notably, in the former studies, the vaccine orally administered to guinea pigs was a combination of live-attenuated SF2a and SF3a strains (30) eliciting a different immune response of mostly secretory IgA and a much weaker serum IgG antibody response compared to that induced by the parenteral conjugate vaccines. Moreover, results of another pioneering study, though not measuring specifically cross-reactivity with SF6 O-SPs, showed that either oral immunization or challenge with SF2a bacteria induced cross-reactive antibodies, which recognized more subsets of heterologous serotypes in humans or monkeys than in guinea pigs (59). Also questioning the importance of animal model selection, a recent study underlined some discrepancy between the functional immune response raised in mice and rabbits following administration of GMMA. In agreement with cross-reactivity data, murine SF2a GMMA-induced antisera had strong homologous bactericidal activity albeit a restricted heterologous bactericidal capacity and did not kill SF6 bacteria. Conversely, rabbit SF2a GMMA-induced antisera could kill SF6 bacteria (32).

In conclusion, our results suggest that the SF2a/SF6 cross-reactivity/cross-protection of IgG antibodies induced by SF2a O-SP-based vaccine candidate could lead to an increase in the overall efficacy of a *Shigella* multivalent candidate vaccine. These findings might be of special value in vaccine design, especially when considering infants in LMICs, the primary target population of a *Shigella* vaccine.

## Authors’ contribution

Conceptualization, D.C., V.A., S. M-S., L.A.M. and A.P.; Immunological assays, V.A., S.M-S., A. B., S. M.; Chemical analyses: J.S, G.W.; Data analysis and interpretation, V.A., S.M-S, S.G., D.C, L.A.M., J.S, G.W and A.P; writing—original draft, V.A., S. M-S., D.C.; writing—review and editing, V.A., S. M-S., D.C., L.A.M., A.P, J. S. and G. W.; funding acquisition, D.C.; Supervision, D.C. All the authors reviewed and approved the final version of the manuscript.

## Ethical approval

The study protocol (#2015-060) including exploratory immunological endpoints was reviewed and approved by the Tel Aviv Sourasky Medical Center Institutional Ethics Committee, Institut Pasteur Institutional Review Board, and the Israeli Ministry of Health.

All participants provided written informed consent before enrolment.

## Data Availability Statement

Comprehensive data are currently included in the article Tables & Figures and Supplementary Material. Any additional data supporting the findings of this study without participant personal identifiers will be made available on request from the corresponding author. This manuscript will be further submitted to a journal that has an open access format and offers the CC-BY license.

