## Supplemental Table S1 for "Serum IgG antibodies induced by the synthetic carbohydrate-based conjugate vaccine candidate SF2a-TT15 against *Shigella flexneri* 2a cross-react with the heterologous lipopolysaccharide of *Shigella flexneri* 6"

### Supplementary Materials

#### Table S1: Serum IgG levels against SF2a LPS, SF6 LPSs, EC147 LPS and Sson LPS 28 days after two injections of SF2a-TT15

| Cohort (dose of SF2a-TT15) | Pre/Post vaccination GMT Number of responders p value GMTR | | LPS | | | | | | |
|---|---|---|---|---|---|---|---|---|---|
| | | | **SF2a LPS** | **lowAc1-SF6 LPS** (Sc544 strain) | **lowAc2-SF6 LPS** (MCDC 2924-71 strain) | **Ac-SF6 LPS** (130316 strain) | **noAc-SF6 LPS** (75922 strain) | **EC147 LPS** (O147:H19:F4ac strain) | **Sson LPS** |
| **Cohort 1 (2 µg)** | **GMT (95%CI) N** | **Pre** | 247 (196-311) N=23 | 1049 (733-1502) N=23 | 2163 (1527-3063) N=23 | 162 (131-200) N=23 | 1220 (825-1804) N=23 | 504 (296-859) N=9 | ND |
| | | **Post** | 1751 (1154-2658) N=23 | 2099 (1339-3290) N=23 | 3200 (2001-5118) N=23 | 200 (135-297) N=23 | 2099 (1304-3378) N=23 | 800 (206-3112) N=9 | ND |
| | **Pre/Post vaccination** | **p value*** | <0.001 | <0.001 | 0.016 | 0.129 | 0.002 | 0.282 | ND |
| | | **GMTR** | 7.1 | 2.0 | 1.5 | 1.2 | 1.7 | 1.6 | ND |
| **Cohort 2 (10 µg)** | **GMT (95%CI) N** | **Pre** | 206 (163-261) N=24 | 755 (554-1030) N=24 | 1510 (1107-2059) N=24 | 112 (80-158) N=24 | 823 (593-1144) N=24 | 476 (347-651) N=24 | 438 (278-690) N=23 |
| | | **Post** | 5382 (3518-8232) N=24 | 2851 (1890-4300) N=24 | 3490 (2388-5099) N=24 | 189 (119-300) N=24 | 2851 (1796-4526) N=24 | 1385 (960-1997) N=24 | 377 (238-597) N=23 |
| | **Pre/Post vaccination** | **p value*** | <0.001 | <0.001 | <0.001 | <0.001 | <0.001 | <0.001 | 0.096 |
| | | **GMTR** | 26.1 | 3.8 | 2.3 | 1.7 | 3.5 | 2.9 | 0.9 |

SF2a LPS, SF6 LPSs and EC147 LPS: Pre: baseline; Post: 28 days after second injection (56 days after first injection) of SF2a-TT15

Sson LPS ("negative control LPS"): Pre: baseline; Post: 56 days after vaccination with SF2a-TT15

GMT: Geometric Mean Titer; CI: confidence interval; GMTR: Geometric Mean Titer Ratio; N: number of sera tested; ND: Not determined

*p value from paired t-test
